# Seroprevalence of COVID-19 infection in a densely populated district in the eastern Democratic Republic of Congo

**DOI:** 10.1101/2022.12.21.22283773

**Authors:** Leonid M. Irenge, Homer M. Bulakali, Arthur Irenge Akonkwa, Jérôme Ambroise, Jean-Luc Gala

**Author notes:** Jérôme Ambroise. Jean-Luc Gala^*^.

## Abstract

Data on coronavirus disease 2019 (COVID-19) prevalence in the Democratic Republic of Congo (DRC) are scarce. We conducted a cross-sectional study to determine the seroprevalence of antibodies against anti-severe acute respiratory syndrome coronavirus 2 (SARS-CoV-2) in the slum of Kadutu, city of Bukavu, between June and September 2021. The survey participants were all unvaccinated against SARS-CoV-2. The crude seroprevalence rate was adjusted to the known characteristics of the assay. Participants aged 15 to 49 years old made up 80 % of the population enrolled in the study (n=507; 319 women and 188 men). The overall crude and adjusted seroprevalence rates of antibodies for COVID-19 were 59.7 % (95 % CI 55.4 % - 63.9 %) and 84.0 % (95 % CI 76.2 % to 92.4 %), respectively. This seroprevalence rate indicates widespread dissemination of SARS-CoV-2 in these communities. COVID-19 symptoms were either absent or mild in more than half of the participants with antibodies for COVID-19 and none of the participants with antibodies for COVID-19 required hospitalization. These results suggest that SARS-CoV-2 spread did not appear to be associated with severe symptoms in the population of these settlements and that many cases went unreported in these densely populated locations. The relevance of vaccination in these communities should be thoroughly investigated.

## Introduction

Severe Acute Respiratory Syndrome Coronavirus-2 (SARS-CoV-2), an RNA virus beta-CoV of group 2B virus, is the causative agent of the current Coronavirus Disease 2019 (COVID-19) pandemic, which began in China at the end of 2019 before spreading throughout the world. The disease’s clinical manifestations range from asymptomatic to acute respiratory distress syndrome, which can be fatal [1]. Because of the emergence of SARS-CoV-2 variants linked to successive waves of the COVID-19 disease, SARS-CoV-2 has evolved at a rapid rate since its initial characterization [2]. Furthermore, a long-term COVID-19 disease with sequelae can develop as a result of an acute clinical form [3, 4].

As of November 23^rd^, 2022, there were 639,172,272 confirmed cases of COVID-19 worldwide, with 6458487 deaths [5], with the majority of cases reported in Europe, North America, Latin America, and Asia [6]. The pandemic has not spared the African continent, with 12,114,735 COVID-19 confirmed cases and 256,094 deaths reported as of November 17^th^, 2022 [7]. South Africa and the Maghreb countries have recorded the highest number of cases and deaths due to the COVID-19 pandemic [7, 8]. In contrast, the low number of declared COVID-19 cases in Sub-Saharan African countries tends to suggest that these countries have successfully limited the spread of this pandemic, despite a low vaccination rate against SARS-CoV-2, with only 22% of the continent’s population fully vaccinated [9]. This observation is best illustrated by the Democratic Republic of the Congo (DRC). As of November 17^th,^ 2022, this 90-million-person country had reported only 94,021 COVID-19 cases and 1,365 deaths [7]. Taken at face value, these figures indicate that the country has somehow succeeded in mitigating the circulation of SARS-CoV-2, even so, the country has large densely populated cities without improved sanitation facilities and basic infrastructure and wherein applying mitigation measures such as social distancing and good hygiene practices have been difficult to implement. Moreover, DRC has one of the lowest vaccination rates in the world, with less than 4% of the population fully vaccinated [9]. It should be noted that the global rate does not reflect the country’s local vaccinations rates. For instance, the current vaccination rate in the easternmost DRC city of Bukavu is of 1.2% of the population, with major variations between different districts of the town [10]. It has been reported in other developing countries that highly densely populated areas are at risk for high transmission of SARS-CoV-2 [11, 12]. This perplexing situation in the DRC may be caused by a significant underestimation of the total number of cases due to the country’s limited testing capacity [13]. As a result, the current study aimed to assess the level of seroprevalence of antibodies for COVID-19 in the informal densely populated district of Kadutu in the easternmost town of Bukavu in DRC. This district is for the most part made up of informal settlements lacking basic infrastructure and therefore poses a high risk for SARS-CoV-2 transmission, as evidenced by studies in similar settlements in other developing countries [11, 12].

This study’s findings will shed light on the current state of seroprevalence of antibodies for COVID-19 in this overcrowded informal settlement in the DRC.

## Methods

A cross-sectional study was conducted at Berna Clinic between June 24^th^ and September 15^th^, 2021, in the densely populated Kadutu district of the town of Bukavu the DRC. Bukavu, the capital of South Kivu province, has a population of 1,886,947 people living in an area of 45,8 km2 [14]. The city is divided into three districts: Bagira, Ibanda, and Kadutu. The district of Kadutu (619,266 inhabitants on an area of 10,9 km2) is densely populated (56813 inhabitants per km2) [14], with few people (334 out of 619266) vaccinated against SARS-CoV-2 [10]. This district comprises six uneven settlements: Cimpunda (18.8% of the total area), Kalere (53.1%), Kasali (9.0%), Mosala (12.1%), and Nyamugo (5.2%). Each neighbor is further divided in several cells, and each cell is constituted of up to six streets. The sample size (n) was calculated using the formula n = Z^2^p(1-p)/d^2^ [15], based on the following assumptions: i) a confidence limit of 95% corresponding to Z = 1,96, ii) a seroprevalence (p) of 50%, given that earlier studies in the same town pointed to a high prevalence [16, 17], and iii) a desired precision (d) of 5%. The sample size was increased by 30% to compensate for nonresponsive participants, as recommended by Israel [18]. Accordingly, the target sample was set at a minimum of 499 participants. Estimating an average of five enrolled participants per household, the study planned to enroll a minimum 100 households. In order to reach this threshold, and assuming that a high proportion of households would be reluctant to enroll in the survey, we set the goal of contacting 130 households in order to request their participation in the survey. Households selection was conducted using multi-stage random sampling. In the first step, the number of households allocated to each of the five settlements was calculated proportionately to its theoretical demographic weight.

In the absence of any detailed population data per settlement, it was assumed that all the 5 settlements had the same population density. Accordingly, 69, 26, 15, 12, and 8 households were selected randomly in Kalere, Cimpunda, Mosala, Kasali, and Nyamugo settlements, respectively. Finally, a reserve of 40 households was randomly selected proportionally to the demographic weight of the settlements of Kalere, Cimpunda, and Mosala (25, 9, and 6, households respectively), and used to replace households which declined participation in the survey, or to compensate for absentees within households at the time of sampling (Figure 1).

**Figure 1.**
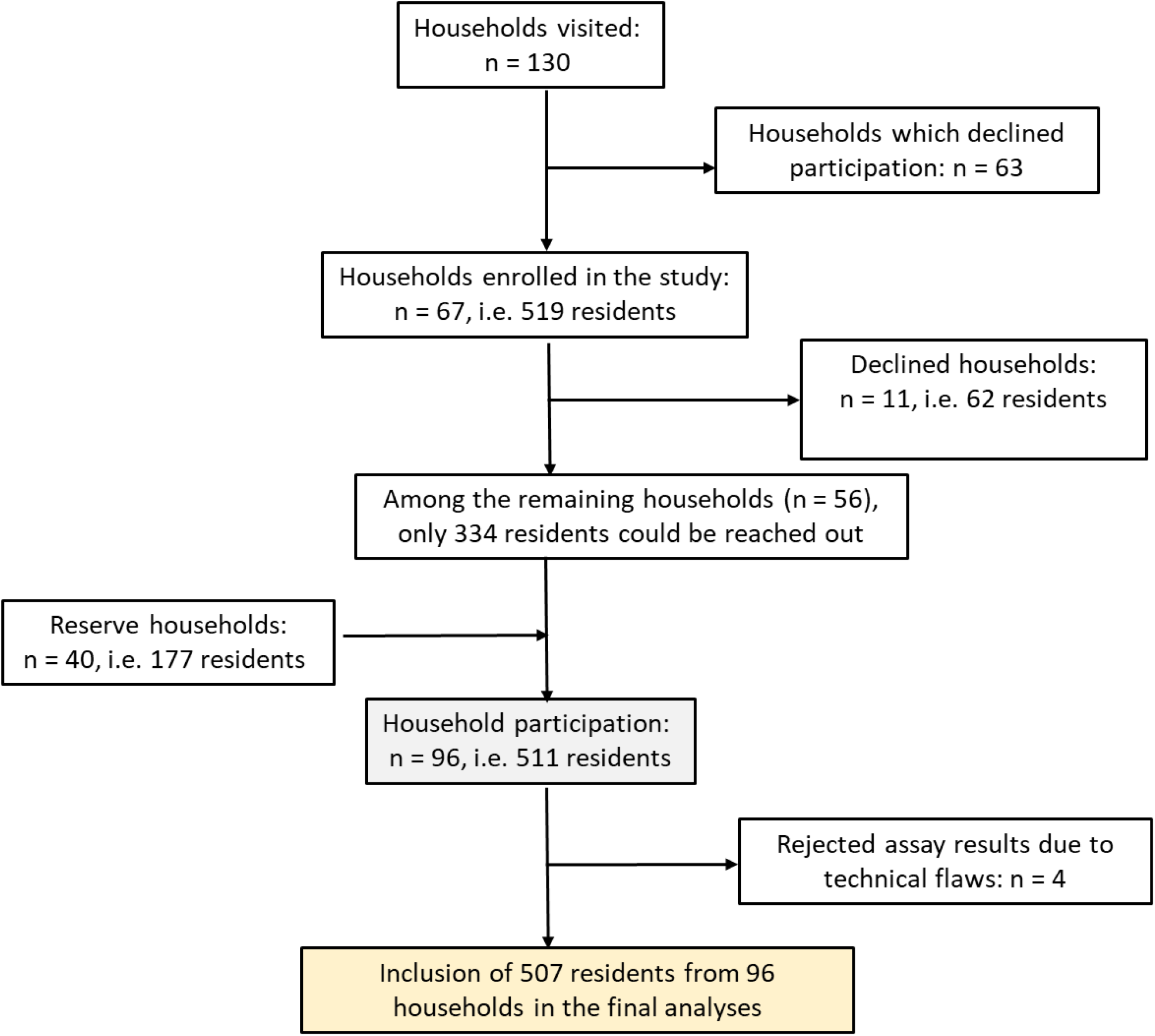
Study enrolment flow chart for the SARS-CoV-2 antibody prevalence survey in Kadutu district, Bukavu, DRC.

### Data collection

During the preliminary phase, a team of four healthcare workers from the Berna Clinic in the Kadutu district went door to door to inform the selected household members about the survey’s objectives of the study. The inclusion criteria were i) the continuous presence of the household in the neighborhood since February 14^th^, 2020 the day when the first COVID-19 case was recorded on the African continent [19], and ii) the lack of any record of vaccination against SARS-CoV-2. All selected participants were interviewed for sociodemographic data such as age and gender, and they completed a questionnaire about whether they had experienced COVID-19 symptoms in the previous 6 months (Table 1).

**Table 1.** Questionnaire answered by people surveyed.

|  | Yes | No |
| --- | --- | --- |
| Have you developed any of the following symptoms in the last 6 months? |  |  |
| -Fever |  |  |
| -Coughing |  |  |
| -Myalgia |  |  |
| -Headaches |  |  |
| -Dysgeusia |  |  |
| -Anosmia |  |  |
| -Respiratory distress |  |  |

### Serological testing for detection of antibodies for COVID-19

The QuickZen® COVID-19 IgM/IgG kit (ZenTech, Angleur, Belgium), an immune colloidal gold test kit, was used to detect IgM and IgG against SARS-CoV-2 S-RBD (receptor-binding domain of the S protein). Fresh finger-prick blood was drawn from each participant and dropped directly into the well of the QuickZen® lateral flow. The results were interpreted following the manufacturer’s recommendations. A positive result was defined as the presence of both the sample (IgG and/or IgM) and control band of the test.

The crude seroprevalence (proportion of participants positive for antibodies against COVID-19) was adjusted using the standard correction formula based on Bayes rule (i.e., adjusted prevalence = (crude prevalence + specificity-1) / sensitivity + specificity – 1) [20, 21], while taking into account the sensitivity (71.1%) and the specificity (100%) of the QuickZen® assay, as determined by Montesinos et al [22]. In addition, the QuickZen® assay was also tested on 49 pre-COVID-19 serum samples which were collected in the district of Kadutu between April 2004 and May 2005.Statistical analysis.

### R version 4.1.2 was used to conduct all statistical analyses

Chi-square association tests were used to compare the gender and age distributions of people who had antibodies for COVID-19 versus those who did not. Chi-square association tests were also used to assess the potential link between antibodies for COVID-19status and a variety of symptoms reported in the six months preceding serological testing. A p-value <0.05 was considered statistically significant.

### Ethical considerations

The Université Catholique de Bukavu’s Internal Review Board (UCB/CIES/NC/02312021) reviewed and approved this study.

Before enrollment, all participants provided written consent or assent. Informed consent was obtained from the children’s parents or guardians. On behalf of their children, a parent or his representative signed the informed consent form. Informed consent was also obtained from each of the participants whose pre-COVID-19 serum samples were used for testing for antibodies using the Quickzen® kit. To allow for anonymous analysis, the identities of participants were removed from the samples.

## Results

The study enrolled a total of 511 people (322 women and 189 men) from 98 households. However, assays from 4 participants were rejected, bringing the total of participants to 507 (319 women and 188 men). Table 2 shows the demographic characteristics of those surveyed. In total, 303 tested positive, resulting in an overall crude seroprevalence of antibodies for COVID-19 of 59.7% (95% CI: 55.4% - 63.9%). The adjusted seroprevalence of antibodies for COVID-19 was 84.0% (95% CI: 76.2% - 92.4%). In the 303 positive tests, the antibody distribution was as follows: IgM band only 26.4% (n = 80), IgG band only 43.9% (n = 133), and both IgM and IgG bands 29.7% (n = 90).

**Table 2.**
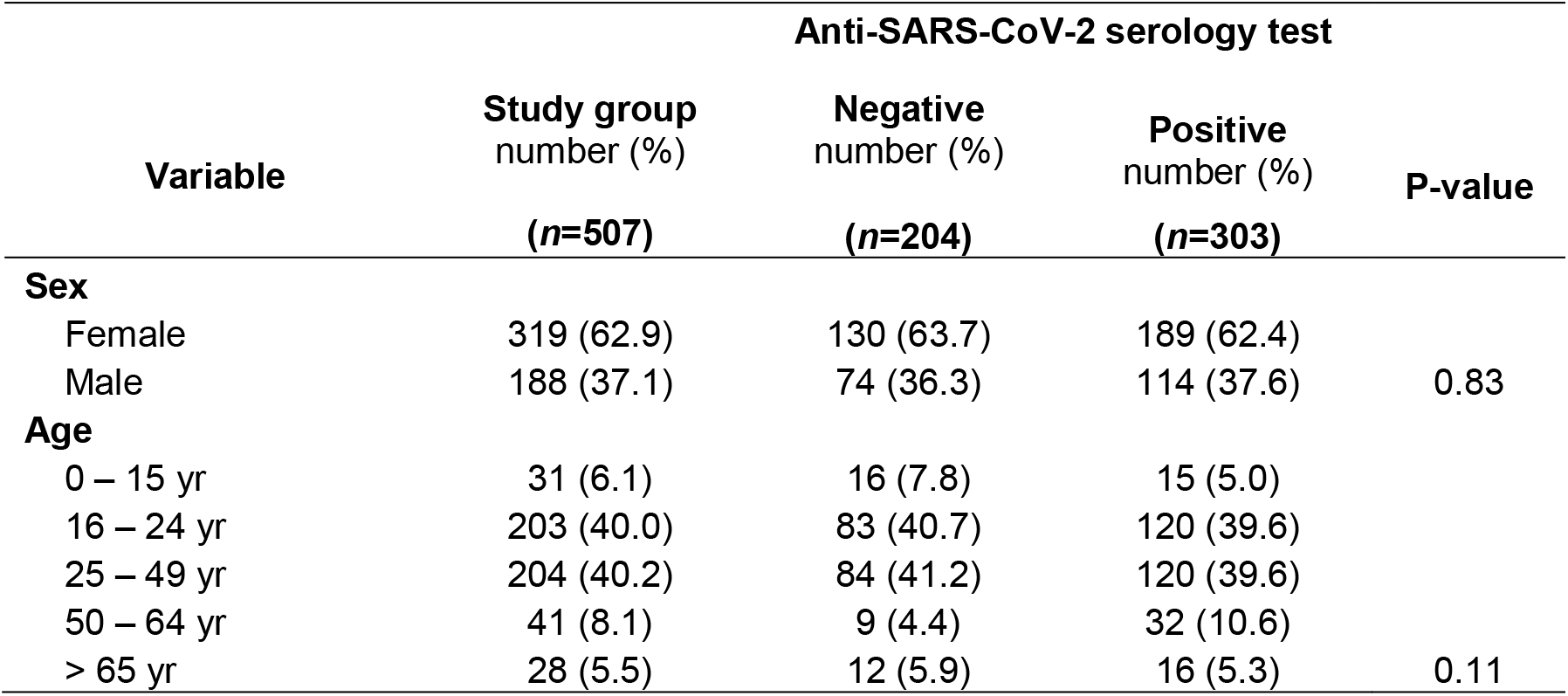
Sex and age distribution as observed in all participants and each study group.

There was no significant relationship between anti-SARS-CoV-2 antibody status and sex (p = 0.83), as evidenced by the very similar sex distribution of people with and without antibodies for COVID-19. Similarly, no significant relationship between antibodies for COVID-19status and age distribution were found (p = 0.11).

Noticeably, none of 49 pre-COVID-19 serum samples assayed was tested positive for anti-SARS-CoV-2 antibodies.

Table 3 shows the proportions of participants with common COVID-19 symptoms. All of the symptoms reported by participants in the six months before the serological testing were minor. No one who tested positive for antibodies for COVID-19 needed to be hospitalized. In participants with (n=303) and without (n=204) antibodies for COVID-19, no symptoms were recalled in 180 (59.4%) and 110 (53.9%) participants, respectively. There was no significant relationship between antibodies for COVID-19 status and any of the symptoms reported.

**Table 3.**
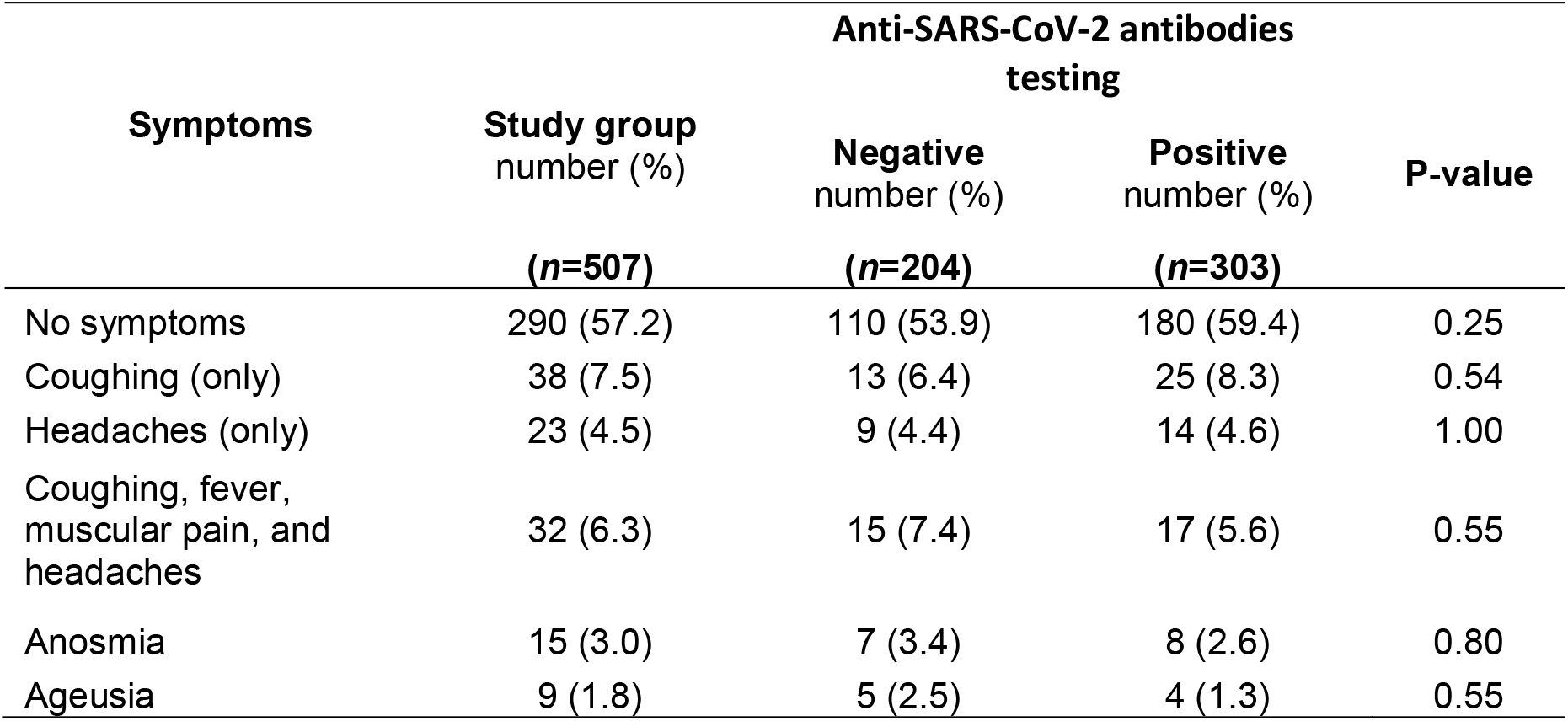
Proportions of participants with symptoms in the study group.

## Discussion

We present the results of a population-based seroprevalence survey conducted in the densely populated informal settlement of Kadutu in the easternmost town of Bukavu, (DRC). We found a crude and adjusted seroprevalence for antibodies for COVID-19 of 59.7 % and 84.0 %, respectively, indicating a very high SARS-CoV-2 transmission rate in this overcrowded urban settlement where vaccination against SARS-CoV-2 has struggled to take off, as highlighted by the current vaccination rate of 0.8 % [10] far below the national average, though higher than the vaccination rate of 0.05% by September 15^th^, 2021 (i.e., the end of this study) [10]. This high transmission rate is most likely related to difficulties in implementing mitigation measures that have been used globally (i.e. social distancing and good hygiene practices). Our data suggest that COVID-19 cases are significantly underreported in the nation. Given the ineffective handling of the COVID-19 pandemic in the DRC, this is not at all surprising [13]. In such cases, seroprevalence studies of COVID-19 antibodies in unvaccinated populations may be able to shed light on the country’s current SARS-CoV-2 transmission. Sadly, there are few seroprevalence survey results in unvaccinated populations in the DRC. A survey conducted during the first COVID-19 wave in Kinshasa metropolis, a DRC city with a population of more than 12 million people, revealed a seroprevalence of antibodies for COVID-19 of 16.6% [23]. In another serosurvey conducted during the first epidemic COVID-19 wave, healthcare workers at the Panzi hospital (Bukavu, DRC) had a seroprevalence antibodies for COVID-19 of 41.2 % [16]. Our findings are consistent with those found in other overcrowded settlements in developing countries [11, 12].

The absence of participants with severe symptoms, which could have necessitated hospitalization, is a startling observation. Our findings are consistent with what has previously been reported in similar settings. Factors that could explain Africa’s low morbidity and mortality rates have been discussed elsewhere [24]. Other potential factors that could contribute to COVID-19’s low morbidity and mortality in Africa include the presence of cross-reacting antibodies to other coronaviruses that have not been thoroughly studied [24, 25] or to other diseases such as malaria [26]. It can be hypothesised that the population, due to the number of various recurrent infectious diseases to which they are chronically exposed, had the capacity to rapidly acquire effective protective immunity against severe forms of COVID-19, stimulated by the high circulation of SARS-CoV-2. The relatively young age of the study participants, 80% of whom were under 49 years of age, is consistent with this hypothesis, which would explain the low demand for medical care and the low mortality rate from COVID-19 compared to other continents.

This study does have some limitations. The population from Kasali and Nyamugo, two settlements far from the Berna Clinic, was the least likely to participate in the survey. This could be attributed to their weak interaction with the Berna Clinic which is located in the Cimpunda settlement. Accordingly, households of settlements close to the Berna clinic (Cimpunda, Kalere, and Mosala) were overrepresented compared to those from Kasali and Nyamugo settlements. This sampling process might have led to selection bias which could have impacted the prevalence estimation. Accordingly, projecting our survey results to the entire district and the entire town, as well as to similar communities across the country is not recommended. To circumvent the limitations related to potentially imperfect positive and negative predictive values of the QuickZen® test, we have reported both the crude and the adjusted seroprevalence rates with confidence intervals, while also considering the specificity and sensitivity of the test. Another limitation is that anti-SARS-CoV-2 antibodies are detected using a qualitative test, which cannot reveal the quantitative value or rate of growth in anti-SARS-CoV-2 antibody levels. This limits the breadth of measuring the population’s immunological state, because a positive result with insufficient antibodies may not protect against SARS-CoV-2 re-infection. Despite these limitations, our data suggest that SARS-CoV-2 has spread widely in this crowded Kadutu district and, most likely, in analogous areas across the country, but more studies are needed to confirm this assumption. The overall high ratio of IgM (170 / 303, 56.1%) among participants with anti-SARS-CoV-2 antibodies is consistent with a recent transmission of the SARS-CoV-2 virus in that district. The low official results may be explained by the country’s insufficient testing capacity for reverse transcription-polymerase chain reaction (RT-PCR) detection [13, 27].

In conclusion, our study found a high seroprevalence of anti-SARS-CoV-2 antibodies in the surveyed people from the overcrowded settlement of Kadutu, indicating a high circulation of the SARS-CoV-2 among its residents. It also shows that the majority of those surveyed either had mild COVID-19 symptoms or no symptoms at all. While our findings cannot be used to infer anti-SARS-CoV-2 seroprevalence for other settlements in the country, a similar trend of high anti-SARS-CoV-2 seroprevalence cannot be ruled out. In order to better understand the dynamics of SARS-CoV-2 transmission in the country, our findings urge more serosurveys to be conducted in rural areas with low population density as well as other urban areas across the nation. Such findings could be used in these communities to disseminate accurate information about COVID-19, which is still widely believed by many to be a hoax designed to induce people to receive unnecessary vaccinations against a non-existent disease. As a first step, our data might be distributed to participants and reputable community leaders. In a subsequent step, these community leaders could make use of these findings to emphasize the significance of putting preventive measures in place with the goal of reducing the spread of COVID-19 in these settlements in order to protect those who are particularly vulnerable, such as the elderly and immunocompromised individuals. These precautions include practical ones like mask use and, most importantly, frequent hand washing. Given the promiscuity in these settlements, social distancing seems difficult to implement to implement.Finally, given the obvious shortcomings of COVID-19 vaccination in DRC, with less than 4% of the population fully vaccinated [9], the relevance of vaccination in these communities should be thoroughly investigated. Targeting vulnerable people or those without anti-SARS-CoV-2 antibodies in overcrowded DRC town settlements for COVID-19 vaccination could represent an alternative to the current stuttering COVID-19 vaccination strategy in DRC.

## Data Availability

The databases used and/or analyzed during the current study are available from the corresponding author upon reasonable request.

## Acknowledgments

We thank the personnel of the Berna Clinic for their technical support in performing public sensitization for the study, for helping participants answer the questionnaire, and for carrying out lateral flow assays for the detection of anti-SARS-CoV-2 antibodies. We acknowledge the assistance of Dr Cubaka M. Jean de Dieu (Programme Elargi de Vaccination/Division Provinciale de la Santé au Sud-Kivu, RD Congo, for providing the data on SARS-CoV-2 vaccination against in Bukavu.

## Data availability statements

The databases used and/or analyzed during the current study are available from the corresponding author upon request.

## Ethical standards

This study was reviewed and approved by the Ethical Review Committee of the Université Catholique de Bukavu (number UCB/CIES/NC/02312021). All participants or their guardians (in the case of children) provided written consent before enrollment.

## References

1. Mehta OP, Bhandari P, Raut A, Kacimi SEO, Huy NT. Coronavirus Disease (COVID-19): Comprehensive Review of Clinical Presentation. Front Public Health. 2020;8:582932.

2. Thakur V, Bhola S, Thakur P, Patel SKS, Kulshrestha S, Ratho RK, et al. Waves and variants of SARS-CoV-2: understanding the causes and effect of the COVID-19 catastrophe. Infection. 2021.

3. Del Rio C, Collins LF, Malani P. Long-term Health Consequences of COVID-19. JAMA. 2020;324(17):1723–4.

4. Mainous AG, 3rd, Rooks BJ, Wu V, Orlando FA. COVID-19 Post-acute Sequelae Among Adults: 12 Month Mortality Risk. Front Med (Lausanne). 2021;8:778434.

5. JHU CfSSaECa. COVID-19 Dashboard 2022 [updated 24-08-2022. Available from: https://coronavirus.jhu.edu/map.html.

6. Assefa Y, Gilks CF, Reid S, van de Pas R, Gete DG, Van Damme W. Analysis of the COVID-19 pandemic: lessons towards a more effective response to public health emergencies. Global Health. 2022;18(1):10.

7. Africa-CDC. Coronavirus Disease 2019 (COVID-19). Latest updates on the COVID-19 crisis from Africa CDC 2022 [updated August 22^th^, 2022. Available from: https://africacdc.org/covid-19/.

8. Tessema SK, Nkengasong JN. Understanding COVID-19 in Africa. Nat Rev Immunol. 2021;21(8):469–70.

9. Coronavirus (COVID-19) Vaccinations 2022 [updated 24-08-22. Available from: https://ourworldindata.org/covid-vaccinations?country=OWID_WRL~COD.

10. Programme Elargi de Vaccination/Division Provinciale de la Santé Sud-Kivu RC. Vaccinations contre le SARS-CoV-2 à Bukavu (RD Congo). 23-08-2022 ed 2022.

11. Ngere I, Dawa J, Hunsperger E, Otieno N, Masika M, Amoth P, et al. High seroprevalence of SARS-CoV-2 but low infection fatality ratio eight months after introduction in Nairobi, Kenya. Int J Infect Dis. 2021;112:25–34.

12. Malani A, Shah D, Kang G, Lobo GN, Shastri J, Mohanan M, et al. Seroprevalence of SARS-CoV-2 in slums versus non-slums in Mumbai, India. Lancet Glob Health. 2021;9(2):e110–e1.

13. Juma CA, Mushabaa NK, Abdu Salam F, Ahmadi A, Lucero-Prisno DE. COVID-19: The Current Situation in the Democratic Republic of Congo. Am J Trop Med Hyg. 2020;103(6):2168–70.

14. Sud-Kivu GdlPd. Tableau synoptique des populations congolaise et étrangère par entité; Rapport annuel 2021. 2021.

15. Pourhoseingholi MA, Vahedi M, Rahimzadeh M. Sample size calculation in medical studies. Gastroenterol Hepatol Bed Bench. 2013;6(1):14–7.

16. Mukwege D, Byabene AK, Akonkwa EM, Dahma H, Dauby N, Cikwanine Buhendwa JP, et al. High SARS-CoV-2 Seroprevalence in Healthcare Workers in Bukavu, Eastern Democratic Republic of Congo. Am J Trop Med Hyg. 2021;104(4):1526–30.

17. Katchunga PB, Murhula A, Akilimali P, Zaluka JC, Karhikalembu R, Makombo M, et al. [Seroprevalence of SARS-CoV-2 antibodies among travellers and workers screened at the Saint Luc Clinic in Bukavu, a city in eastern Democratic Republic of the Congo, from May to August 2020]. Pan Afr Med J. 2021;38:93.

18. Israel GD. Determining Sample Size. University of Florida Cooperative Extension Service, Institute of Food and Agriculture Sciences, EDIS, Florida [Internet]. 1992.

19. Saied AA, Metwally AA, Madkhali NAB, Haque S, Dhama K. Egypt’s COVID-19 Recent Happenings and Perspectives: A Mini-Review. Front Public Health. 2021;9:696082.

20. Sempos CT, Tian L. Adjusting Coronavirus Prevalence Estimates for Laboratory Test Kit Error. Am J Epidemiol. 2021;190(1):109–15.

21. Meyer MJ, Yan S, Schlageter S, Kraemer JD, Rosenberg ES, Stoto MA. Adjusting COVID-19 Seroprevalence Survey Results to Account for Test Sensitivity and Specificity. Am J Epidemiol. 2022;191(4):681–8.

22. Montesinos I, Gruson D, Kabamba B, Dahma H, Van den Wijngaert S, Reza S, et al. Evaluation of two automated and three rapid lateral flow immunoassays for the detection of anti-SARS-CoV-2 antibodies. J Clin Virol. 2020;128:104413.

23. Nkuba AN, Makiala SM, Guichet E, Tshiminyi PM, Bazitama YM, Yambayamba MK, et al. High Prevalence of Anti-Severe Acute Respiratory Syndrome Coronavirus 2 (Anti-SARS-CoV-2) Antibodies After the First Wave of Coronavirus Disease 2019 (COVID-19) in Kinshasa, Democratic Republic of the Congo: Results of a Cross-sectional Household-Based Survey. Clin Infect Dis. 2022;74(5):882–90.

24. Njenga MK, Dawa J, Nanyingi M, Gachohi J, Ngere I, Letko M, et al. Why is There Low Morbidity and Mortality of COVID-19 in Africa? Am J Trop Med Hyg. 2020;103(2):564–9.

25. Tso FY, Lidenge SJ, Pena PB, Clegg AA, Ngowi JR, Mwaiselage J, et al. High prevalence of pre-existing serological cross-reactivity against severe acute respiratory syndrome coronavirus-2 (SARS-CoV-2) in sub-Saharan Africa. Int J Infect Dis. 2021;102:577–83.

26. Arshad AR, Bashir I, Ijaz F, Loh N, Shukla S, Rehman UU, et al. Is COVID-19 Fatality Rate Associated with Malaria Endemicity? Discoveries (Craiova). 2020;8(4):e120.

27. Makulo JR, Mandina MN, Mbala PK, Wumba RD, Akilimali PZ, Nlandu YM, et al. SARS-CoV2 infection in symptomatic patients: interest of serological tests and predictors of mortality: experience of DR Congo. BMC Infect Dis. 2022;22(1):21.

